# Gender Balance and Readability of COVID-19 Scientific Publishing: A Quantitative Analysis of 90,000 Preprint Manuscripts

**DOI:** 10.1101/2021.06.14.21258917

**Authors:** Leo Anthony Celi, Marie-Laure Charpignon, Daniel K. Ebner, Aaron R. Kaufman, Liam G. McCoy, Maria Cecilia Millado, Joel Park, Justin Salciccioli, Julia Situ

## Abstract

Releasing preprints is a popular way to hasten the speed of research but may carry hidden risks for public discourse. The COVID-19 pandemic caused by the novel SARS-CoV-2 infection highlighted the risk of rushing the publication of unvalidated findings, leading to damaging scientific miscommunication in the most extreme scenarios. Several high-profile preprints, later found to be deeply flawed, have indeed exacerbated widespread skepticism about the risks of the COVID-19 disease – at great cost to public health. Here, preprint article quality during the pandemic is examined by distinguishing papers related to COVID-19 from other research studies. Importantly, our analysis also investigated possible factors contributing to manuscript quality by assessing the relationship between preprint quality and gender balance in authorship within each research discipline. Using a comprehensive data set of preprint articles from medRxiv and bioRxiv from January to May 2020, we construct both a new index of manuscript quality including length, readability, and spelling correctness and a measure of gender mix among a manuscript’s authors. We find that papers related to COVID-19 are less well-written than unrelated papers, but that this gap is significantly mitigated by teams with better gender balance, even when controlling for variation by research discipline. Beyond contributing to a systematic evaluation of scientific publishing and dissemination, our results have broader implications for gender and representation as the pandemic has led female researchers to bear more responsibility for childcare under lockdown, inducing additional stress and causing disproportionate harm to women in science.

## Introduction

In peer review settings, newly submitted scientific articles are sent by journals to “peer” academic researchers in the same discipline, subjected to scrutiny, and either accepted, rejected, or returned with constructive commentary to improve the manuscript prior to final acceptance. This process, which has dominated mainstream science for decades^1^, is itself the topic of significant debate within the scientific community. Proponents emphasize peer review as an essential gatekeeper to ensure scientific rigor, while detractors describe the process as excessively slow, persistently fallible, and subject to the idiosyncratic biases of individual reviewers^2^.

The preprint server system seeks to address these concerns by serving as a venue for scientists to share early-stage research, facilitating speed, open collaboration, and public review before submission for peer review^3–7^. Advocates highlight the rapid dissemination of data sets afforded by preprints as well as the sharing of results that would otherwise not be readily available^8,9^. This ecosystem is exemplified in biology by bioRxiv^10^ (founded in 2013) and in medicine by medRxiv^11^ (founded in 2019), inspired by the prominent physical, mathematical, and computer science server arXiv that has existed since 1991^12^. However, the preprint process has attracted its own range of criticisms, with concerns regarding the spread of unverified, inadequately conceived, or poorly executed research^6,13,14^. Findings emanating from research in biology, medicine, and related fields take longer to establish and are more difficult to verify through independent replication because of health data privacy, data acquisition cost, and the duration of lab-based experiments. Notably, many preprints do not make their way to final publication^15^ and the goal of early feedback may be superseded by competing concerns, such as an author’s wish to showcase the novelty of their research and thus establish intellectual provenance^3^.

The rapid unfolding of the COVID-19 pandemic in the Winter and Spring of 2020 placed tremendous strain upon the medical science ecosystem, as investigators hurried to characterize both the novel infection and the impact of the unprecedented public health measures deployed to control it. Despite the best efforts of journal editors and reviewers attempting to expedite the peer review process^16,17^, researchers increasingly turned to preprints^18^. For example, critical findings such as Imperial College’s initial COVID-19 modeling^19^ and the RECOVERY trial’s report on the effectiveness of dexamethasone treatment^20^ were first disseminated via preprint. Simultaneously, papers of questionable quality and veracity, such as a later-withdrawn paper linking sequences in COVID-19 to HIV^21^, also flooded these same preprint platforms, leaving journalists with the difficult task of sorting fact from fiction in communicating with the public. Notably, this retracted paper remains the most downloaded and retweeted bioRxiv/medRxiv preprint of all time^22^, offering a cautionary tale as to the challenges of preprint publishing. The COVID-19 pandemic also placed particular strain upon female academics, with notedly inequitable burdens faced in care activities and adjustment to work-from-home, as well as exacerbation of gender disparity in published scientific work^23–26^.

Our goal was to specifically examine this rapid expansion of preprint papers and investigate concerns related to the writing quality of manuscripts and to the balance in the authors’ gender. We generated a sizable database of 240,181 preprint papers (including all versions of a given manuscript) posted onto bioRxiv and medRxiv in the Winter and Spring of 2020 and performed quantitative analyses of the content of these works. We assessed markers of manuscript quality, including length, readability, and spelling correctness, as well as markers of gender representation including the gender balance ratio of authorship. We also examined interactions between these factors, with the aim of understanding the connection between research discipline and gender mix in authorship and metrics of manuscript quality.

## Materials and Methods

Initially, the 91,056 unique preprint documents published to medRxiv (6.9%, n=6,291) or bioRxiv (93.1%, n=84,644) between January 1, 2020 and June 3, 2020 were compiled using the ROpenSci library. To remove redundancies, only the first version of a given manuscript was considered in cases where several were posted online. The medRxiv Application Programming Interface (API) identified 11,047 (i.e., 12.1%) of these articles, 8,665 from medRxiv (i.e., 56.0%) and 2,383 (i.e., 1.2%) from bioRxiv, as being COVID-19-related. Whether or not a paper was COVID-19-related was included as an independent binary variable in the analysis. Furthermore, the API provided additional data including article metadata (e.g., DOI, title, date published, article link, pdf link), authors, article categories (e.g., genetics and genomic medicine, hematology, epidemiology), and affiliated institutions. Using the requests library from Python, the corresponding PDF files were downloaded. These were then parsed using the PdfFileReader function from the PyPDF2 Python package to extract text from the preprint articles. Further, the pyspellchecker Python package was used to quantify the rate of spelling mistakes in the article text^27^.

Two variables, preprint text readability and gender balance in authorship, were then calculated as proxy measures of manuscript quality. Of note, the first variable was composite; it contains a wide range of measures capturing text readability^28^. Lower readability has indeed been implicated as an indicator of less accessibility to interdisciplinary audiences and to non-experts^29,30^. Readability analysis was conducted using Python packages textstat and readability metrics^31,32^. Metrics used include: the percent of words misspelled, the Flesch reading ease scale, the Flesch-Kincaid Reading Level, the Coleman-Liau Index (CLI), the Automated Readability Index (ARI), the SMOG Index, the Gunning-Fog score, the Linsear Write Formula, the Dale-Chall Readability score, as well as the number of unique words, number of syllables, and number of sentences. The Dale-Chall readability test, which grades text difficulty based on word length, was applied to each article in the data set^33^. Unlike the other indices, both the CLI^34^ and the ARI^35^ rely on the number of characters per word, instead of the more commonly used number of syllables per word. Using these correlated metrics, a composite index was constructed by conducting a principal components analysis^36^ using the R library FactoMineR^37^. Then, we selected the first principal component for each preprint, which accounted for 68.5% of the variance explained. Rather than relying on the idiosyncratic properties of any single readability metric, this first principal component was used as the primary measure of readability.

Next, the gender balance ratio was considered. For each author, an automated process was deployed to infer and assign a gender. Initially, the first names of all authors were extracted from each article using regular expressions in Python. First names were pre-processed to remove hyphens, periods, and replace accented letters with the unaccented versions. The resulting list of first names was subsequently processed through the Genderize.io API^38^. The API contains a collection of previously annotated first names and their reported genders. Given a first name input, the API would then provide the empirical probability of a male or female gender. Based on this empirical probability and a threshold fixed at 0.5, we assigned the most-likely gender corresponding to the first name. When preprints have a single author, the metric of gender balance is not as relevant, since in such cases the percentage of female authors is either 0% or 100%. Of note, preprints in biology, medicine, and related disciplines often include a panel of contributors, as opposed to publications in the social sciences, where single authorship is more prevalent^39^.

Furthermore, we extracted the subject area of each medRxiv and bioRxiv preprint using the “Subject Area” field displayed on the preprint server’s landing page (e.g., “Infectious Diseases (except HIV/AIDS)” for medRxiv, “Scientific Communication and Education” for bioRxiv). This additional preprint characteristic was used to further stratify the set of preprints hosted by each server by subject area and test for heterogeneity of outcomes across research domains.

To analyze temporal differences between papers related to COVID-19 or not, we followed a difference-in-differences strategy^40^. This difference-in-differences allowed us to hold constant factors affecting the overall quality of academic articles and isolate the changes related to COVID-19 articles specifically. The data extraction and analyses were performed using R 4.0.4 and Python 3.9.1.

## Results

Across the two primary variables of interest, namely readability and gender balance in authorship, we found that manuscript quality changed between January and June 2020 among papers related to COVID-19, whereas it remained constant over time among non-COVID-19 papers.

The analysis stratified by preprint server (i.e., studying medRxiv and bioRxiv separately) led to mixed results. In the medRxiv sample, we found that COVID-19 preprints released between February and April 2020 were substantially more difficult to read than preprints unrelated to COVID-19. However, by mid-May 2020, the difference between papers related to COVID-19 and others was negligible. Conversely, COVID-19 papers posted on bioRxiv were *more* readable on average than medRxiv preprints (Figure 1). This difference might be attributed to the preprint servers’ respective disciplines: while medRxiv hosts papers related to public health and the social science of medicine, bioRxiv papers are more rooted in biology and biochemistry.

**Figure 1:**
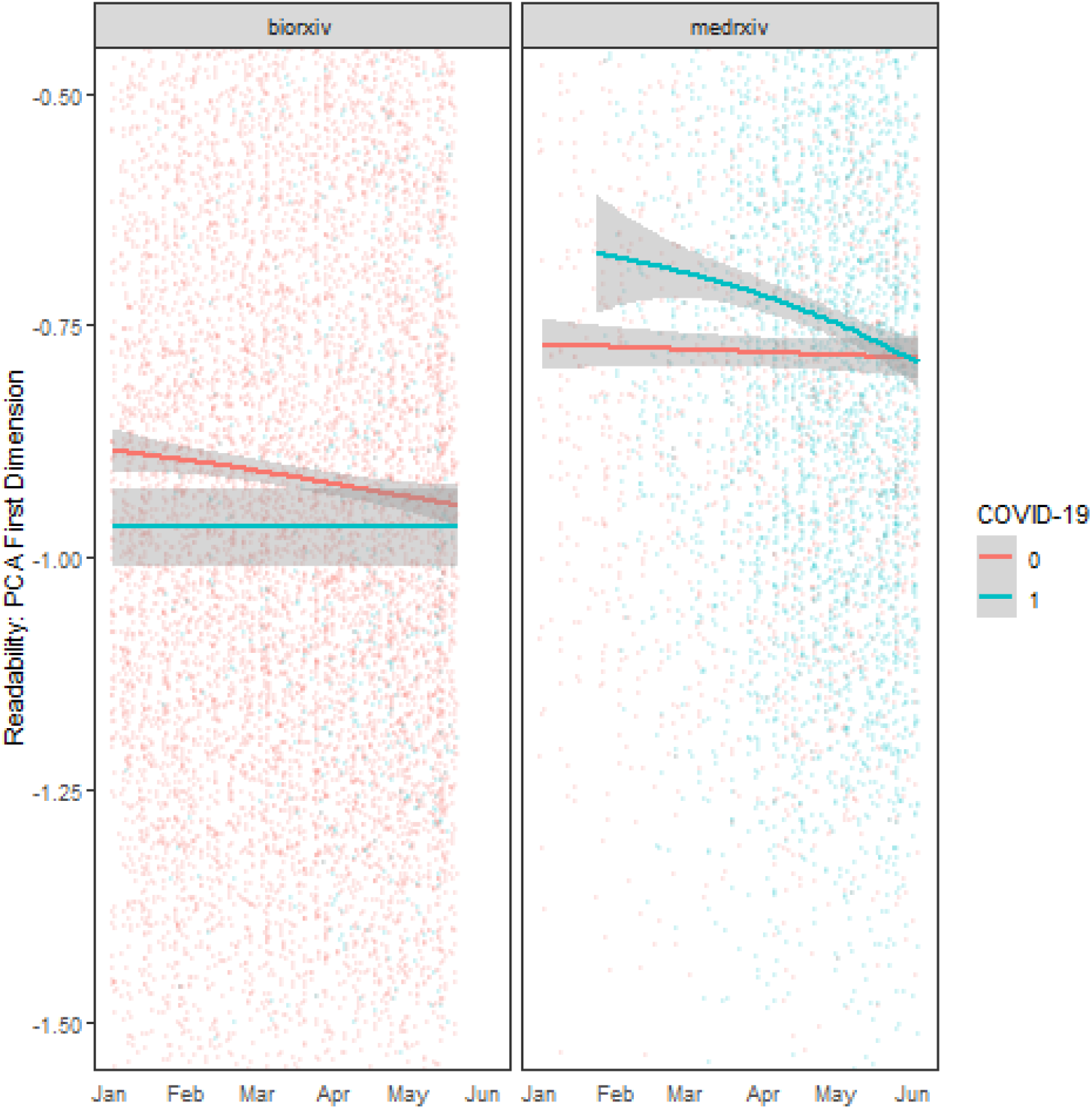
Manuscript readability among bioRxiv and medRxiv preprints over time, between January and June of 2020. On each panel, the y-axis represents the value of the composite readability index corresponding to the first principal component. The lines correspond to daily averages (based on preprint release date, first version) among COVID-19-related papers (blue) and other research studies (pink), while shaded areas represent 95% confidence intervals. Each dot maps to a preprint article posted on either bioRxiv (left panel) or medRxiv (right panel).

In terms of gender balance in authorship, results were consistent across preprint servers. While the proportion of female authors on non-COVID-19 papers remained constant over time (Figure 2, pink line), the proportion of female authors on COVID-19-related papers was substantially lower, except in mid-March 2020. Similar to the time-varying difference in readability scores, the gap between COVID-19-related preprints and others decreased over time and approached zero in June 2020. Overall, both medRxiv and bioRxiv papers related to COVID-19 had substantially fewer female authors than other papers they host; these results held when controlling for variation by research discipline. For example, Figure 3 illustrates the case of epidemiology and infectious diseases (for medRxiv and bioRxiv servers in aggregate), two research sub-disciplines predominantly featured among preprints released on both medRxiv and bioRxiv from January to June 2020. The variation in gender balance in authorship associated with a paper’s relevance to COVID-19 aligns with the underlying heterogeneity in gender balance in authorship observed among research sub-disciplines (e.g., immunology-related preprints generally have a better gender mix than epidemiology preprints; see Appendix Table 1). Developmental biology, genetic medicine, and psychiatry were the research sub-disciplines with the best gender balance ratios, while bioinformatics, paleontology, and biophysics presented the opposite pattern. Among COVID-19 preprints, the fields of public health and infectious diseases had more gender-balanced groups of authors, while epidemiology had less balanced teams. A large proportion of preprints was written by male-only (24.9%, n=22,603) or female-only teams (4.3%, n=3,953), with similar results for medRxiv (male-only: 23.3%, n=1,465; female-only: 2.4%, n=154) and bioRxiv (male-only: 25.0%, n=21,138; female-only: 4.5%, n=3,799). This pattern was more pronounced for COVID-19-related (male-only: 29.4%, n=870; female-only: 2.6%, n=76) than for other preprints (male-only: 24.7%, n=21,733; female-only: 4.4%, n=3877). Of note, only 3.2% (n=2,868) of the considered preprints were single-authored (n=2,481 or 2.9% of bioRxiv and n=382 or 6.2% of medRxiv preprints, respectively). Among single-authored preprints, 13.2% (n=349) were written by female researchers, predominantly in the fields of bioinformatics, neuroscience, and evolutionary biology. Refer to Appendix Table 1 for more details.

**Figure 2:**
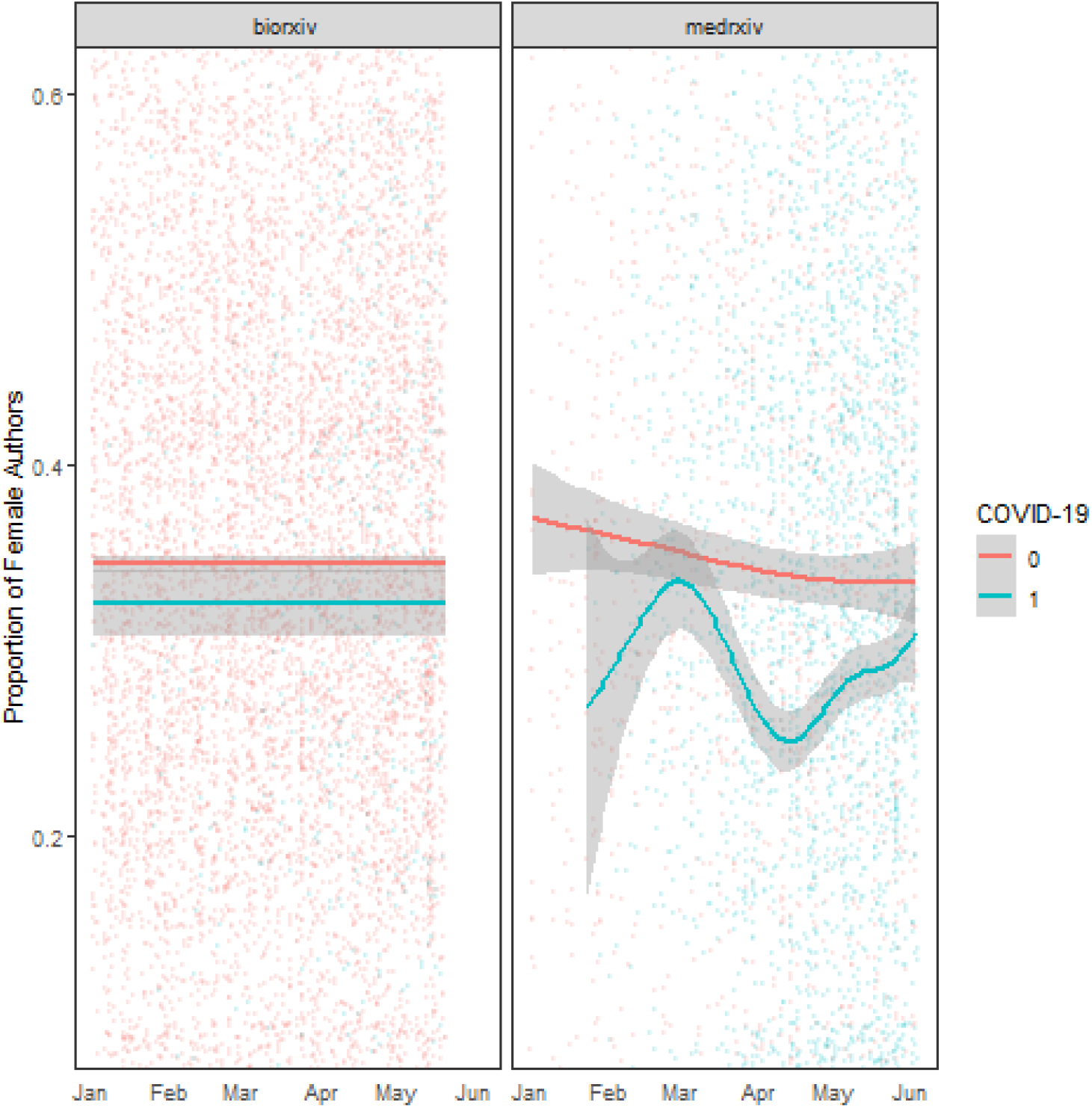
Female authorship among bioRxiv and medRxiv preprints over time, between January and June of 2020. On each panel, the y-axis represents the proportion of female authors. The lines correspond to daily averages (based on preprint release date, first version) among COVID-19-related papers (blue) and other research studies (pink), while shaded areas represent 95% confidence intervals. Each dot maps to a preprint article posted on either bioRxiv (left panel) or medRxiv (right panel).

**Figure 3:**
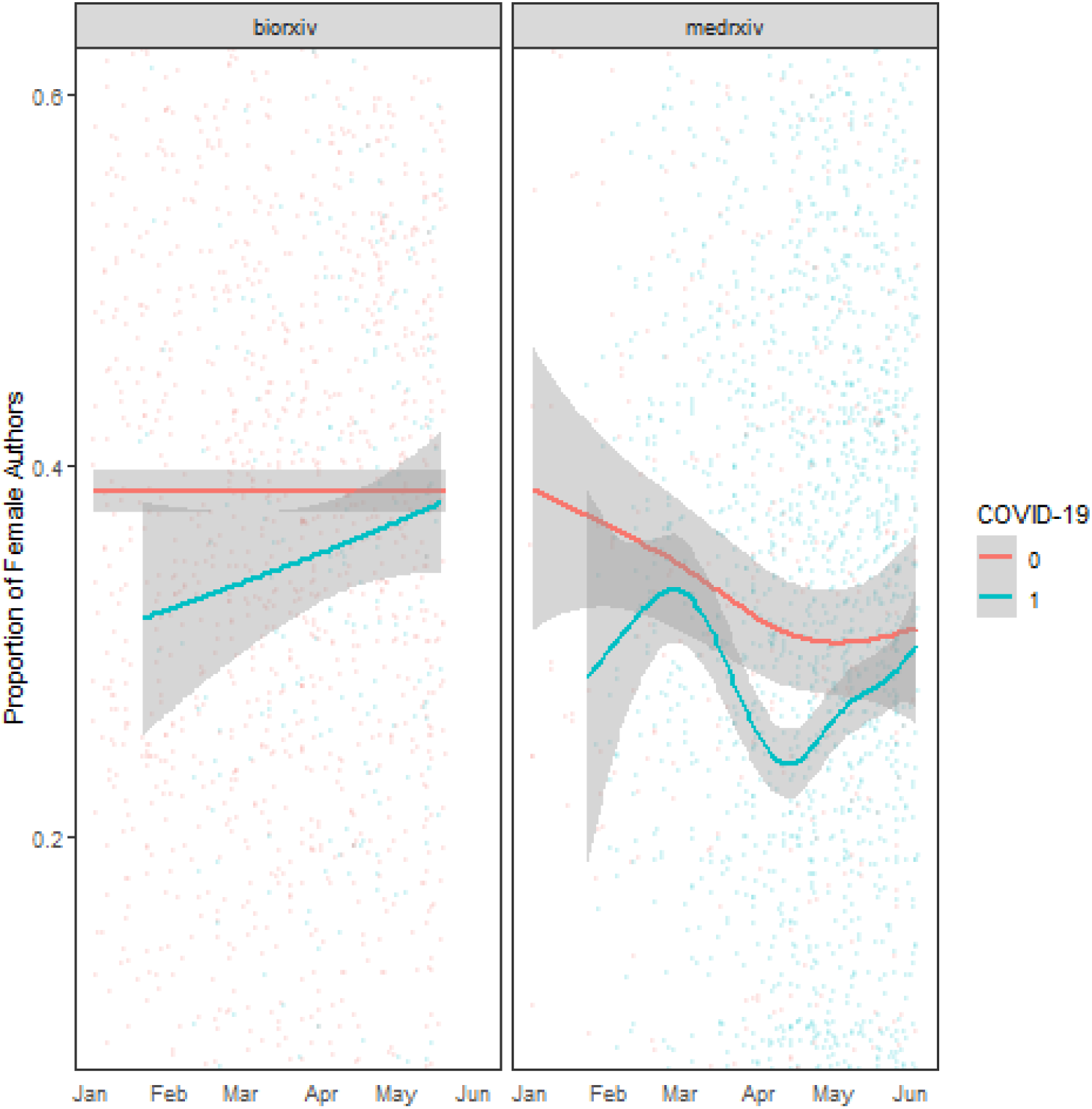
Female authorship among among bioRxiv preprints related to immunology, pathology, and microbiology and medRxiv preprints related to epidemiology and infectious diseases over time, between January and June of 2020. On each panel, the y-axis represents the proportion of female authors. The lines correspond to daily averages (based on preprint release date, first version) among COVID-19-related papers (blue) and other research studies (pink), while shaded areas represent 95% confidence intervals. Each dot maps to a preprint article posted on either bioRxiv (left panel) or medRxiv (right panel).

Differences in gender balance in authorship between preprints might have substantial implications in terms of science communication during the pandemic. For example, we found that more gender-balanced author teams produced more readable preprints. The preprint quality gap between COVID-19 and non-COVID-19 papers was the highest for all-male teams. Notably, this gap shrank as the gender composition of the team approached or exceeded parity (Figure 4).

**Figure 4:**
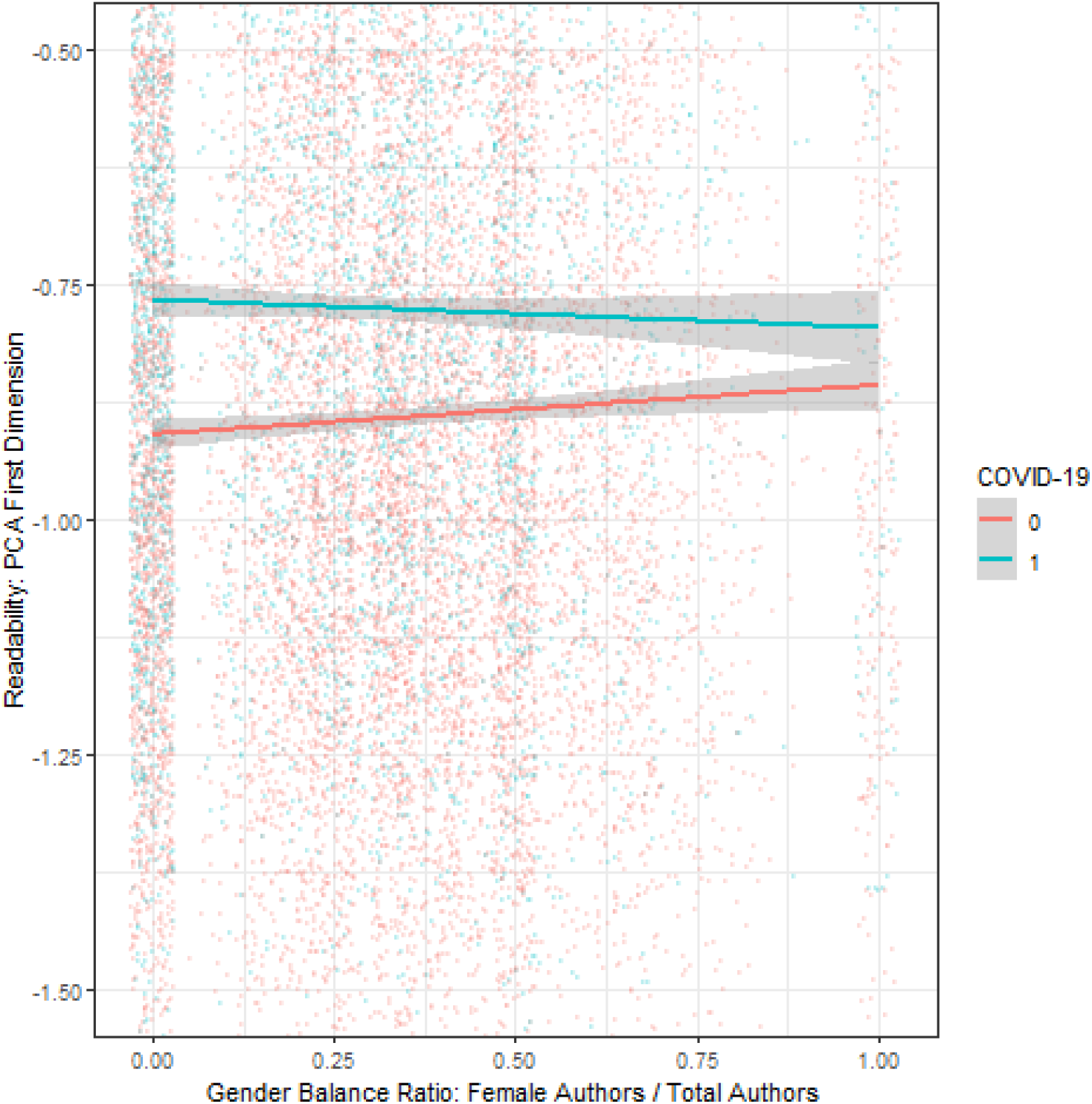
Manuscript readability as a function of gender balance in authorship among preprints posted between January and June of 2020. The x-axis represents the proportion of female authors (from 0% to 100%). The y-axis represents the value of the composite readability index corresponding to the first principal component. The lines correspond to averages among COVID-19-related papers (blue) and other research studies (pink), while shaded areas represent 95% confidence intervals. Each dot maps to a preprint article posted on either bioRxiv or medRxiv (combined into one set).

To further investigate the relationship between gender balance in authorship, manuscript subject area, and readability, we performed four regressions. The first regression considered manuscript readability as a function of gender balance in authorship and whether the paper was related to COVID-19 or not. The second regression included an additional interaction factor between these two variables. Models 3 and 4 were similar to models 1 and 2 respectively, but with the addition of fixed effects by research discipline, as determined by subject area tags used by the preprint servers. The corresponding results are presented in Table 1 below.

**Table 1:** Multivariate regression results. The dependent variable is Readability: PCA First Dimension. Regressors include the gender balance ratio (continuous, between 0% and 100%), the COVID-19 binary variable, the interaction of these two variables, as well as sub-discipline fixed effects (x dummy variables and one reference). Four models were evaluated. Adjustments for the gender balance ratio and whether preprints were COVID-19-related or not were made in all models. Two models considered the interaction of these two variables. Two models included sub-discipline fixed effects. An intercept was considered in all models.

|  | *Dependent variable:* |  |  |  |
| --- | --- | --- | --- | --- |
|  | Readability |  |  |  |
|  | (1) | (2) | (3) | (4) |
| Gender Balance Ratio | 0.032**<br>(0.016) | 0.052***<br>(0.019) | 0.040**<br>(0.016) | 0.054***<br>(0.019) |
| COVID-19 | 0.118***<br>(0.009) | 0.143***<br>(0.014) | −0.024*<br>(0.013) | −0.007<br>(0.017) |
| Interaction |  | −0.082**<br>(0.037) |  | −0.055<br>(0.037) |
| Sub-discipline Fixed Effects-medicine | No | No | Yes<br>(0.492) | Yes<br>(0.492) |
| Constant | −0.902***<br>(0.007) | −0.909***<br>(0.008) | −1.163**<br>(0.474) | −1.164**<br>(0.474) |
| Observations | 15,764 | 15,764 | 15,764 | 15,764 |
| R<sup>2</sup> | 0.011 | 0.012 | 0.042 | 0.042 |
| Adjusted R<sup>2</sup> | 0.011 | 0.011 | 0.038 | 0.038 |

*Note:*
\*p<0.1; \*\*p<0.05; \*\*\*p<0.01

In all four models, the positive regression coefficient for the gender balance ratio indicates that this variable is an important positive predictor of manuscript readability in general, irrespective of whether the preprint is COVID-19-related or not (Table 1, coefficients for the “Gender Balance Ratio” variable). For interpretation, models 3 and 4 are favored over models 1 and 2 because of their increased robustness. When controlling for research sub-discipline via fixed effects (models 3 and 4), the regression coefficient for the COVID-19 binary variable is negative, suggesting that COVID-19-related preprints are less readable than others (Table 1, coefficients for the “COVID-19” variable). The negative coefficient for the interaction term in models 2 and 4 (Table 1, coefficients for the “Interaction” term) means that the relationship between gender balance in authorship and preprint readability is less pronounced for COVID-19-related papers than for other preprints released during the same time period. In other words, as the gender balance ratio approaches parity, COVID-19-related papers become more legible, with readability scores getting closer to those of manuscripts released during the same time frame but unrelated to COVID-19.

## Discussion

Prior work has raised concerns about the quality of research methodologies used during the COVID-19 pandemic^6,41–44^. Our results lend credence to these concerns, highlighting the measurable differences between the readability of preprints prior to and during the rapid COVID-19 preprint expansion. Specifically, we found that COVID-19-related medRxiv preprints released between March and May 2020 were less readable than other papers posted during the same period. Importantly, while we demonstrated that gender mix in authorship was positively associated with higher scores for manuscript readability, these three months were marked by a lower proportion of female authors among COVID-19-related preprints.

Our findings contribute to broader work on the topic of readability in scientific research. Readability has received more attention during the COVID-19 pandemic because of the increased consumption of scientific literature by the general public^8,45,46^. In addition, the development of automated metrics providing a well-validated proxy for manuscript readability has facilitated the computational evaluation of the accessibility of scientific work^28^. Given the rise of the Internet as a primary tool for scientific information and dissemination, the combination of poor readability of online medical information and potential scientific errors erodes public trust^47–49^. The COVID-19 pandemic has increased the role of accessibility, with preprint papers being released at a rapid pace and used to guide both individual action and public policy^50^.

In line with earlier research highlighting the impact of COVID-19 on gender equity in medical science^26,51,52^, we found that both medRxiv and bioRxiv preprints related to COVID-19 had fewer female authors than other papers, even after adjusting for variation by research discipline. With the advent of “work-from-home” policies following strict lockdowns, female academics faced significantly increased burdens of childcare relative to their male colleagues^53–57^. Moreover, the pandemic may have deepened sexist biases regarding who is considered an authoritative source of scientific knowledge^53^. Additional contributing factors may include internalized and structural biases in scientific self-assessment, with gender differences in confidence^58–60^, self-perception^61^, and self-promotion well documented in previous studies of academic research environments^62^. Such research work indeed suggests that male authors might be more inclined (either through appropriate confidence or overconfidence) to engage in novel research areas than women with similar levels of expertise, perhaps due to concerns of imposter syndrome^63–65^.

Recognizing and addressing gender diversity challenges in scientific research is critical both to social justice and to the quality of the scientific enterprise. Our findings suggest that the lack of gender balance is detrimental to the quality of research outputs, with a significant decrease in readability observed as the gender balance ratio in authorship diverges from parity. This is in line with prior research demonstrating that gender mix leads to more effective teams^66–68^, better science, and improved patient outcomes^69^. Another possible explanation is that the investigators who strive for gender balance in authorship may be particularly diligent scientists and better research communicators than those who do not cultivate such an endeavor. Future work could examine this intriguing hypothesis: first, by assessing whether an author’s disposition to have more gender-balanced teams is consistent over time and throughout their research work, which could testify to their thoughtful intention or personal values; second, by further measuring how such inclination/attitude correlates with grant outcomes, publications, and citations. Notwithstanding, the situation of early COVID-19 preprints highlights and deepens long-standing gender inequities in research opportunities^70,71^; hence more work is needed to address these disparities at all levels of the research pipeline.

The COVID-19 pandemic has also served to highlight a persistent fear about preprints: that false or misleading results may be propagated to the public by journalists either lacking the requisite technical skills to distinguish carefully validated findings from spurious associations or rushing to disseminate preliminary results. Many journals have already had longstanding norms against discussing preprint research with the media^7^, but this taboo largely fell by the wayside in the process of the early COVID-19 pandemic. The burden of being aware of the impact of a preprint publication falls both upon the researchers drafting the work and the journalists sharing it with a broad audience.

The context of the COVID-19 pandemic further demonstrates the importance of meta-research to better understand and characterize both the internal dynamics of the scientific enterprise, and the ways in which science interacts with public action and public policy. With the rapid pace of social media, preprints are not obscure documents limited in readership to a small part of the scientific community^72^ – the release of a preprint is generally associated with a significant increase in attention that carries forward even to the final published article^73,74^. Further work must be done to better characterize these information and attention flows. Monitoring metrics such as readability, which could be used to evaluate the quality of the scientific production, can help guide the research enterprise as well as structural and institutional responses to ensure accurate and effective science communication. Such responses could include gender policy changes supporting female scientists, shown to improve research quality during the pandemic^75^.

Concerns regarding research quality among COVID-19-related preprints have re-ignited a movement toward the improvement of scientific research quality more holistically^76^. As with many other challenges highlighted by COVID-19, concerns about public trust, research quality, and gender diversity in science are far from novel^77^. However, this crisis has underscored the importance of fostering an inclusive, equitable, and trustworthy medical research ecosystem. The COVID-19 pandemic has revealed the insufficiencies of the peer review system in times of such rapidly evolving crisis, but as exemplified by our analyses, it has also revealed insufficiencies in the preprint ecosystem.

## Limitations and Future Work

The gender inference methods used in this paper, as with any methods outside of direct survey and self-identification, are imperfect and likely to have misgendered a subset of the author population. Additionally, the Genderize.io API was unable to assign a gender to every author name. Moreover, the API is not inclusive beyond the gender binary (e.g., intersectional gender and they/them pronouns are not considered). However, as shown in Appendix Figure 1, our results hold even if we impute all indeterminate genders as female. In addition, we are limited to an extent in our ability to infer what readability implies about any specific papers, since we examined papers in aggregate rather than individually. Finally, we have not yet considered the publication rate and time from initial preprint release to publication. At this stage, our analyses of publication outcomes would indeed be limited by right-censoring, in that the “true” outcome is not available for papers which may be published after our time of analysis (in late 2021, 2022, or later, depending on field norms). Preprint manuscripts are typically en route to peer-reviewed publication, but how many actually obtain publication (and how long the process from preprint release to actual publication takes) may be an important measure of their quality, although the latter may be confounded by differential editorial review processing timelines among scientific and medical journals^78–80^. Given that both medRxiv and bioRxiv track publication status of their manuscripts, future work would include the calculation of both a binary outcome indicating whether a preprint had been published as of November 30, 2020 and a continuous outcome characterizing the time from preprint release to publication of the corresponding peer-reviewed article.

## Conclusion

The COVID-19 pandemic has placed extreme pressure upon medical research and the peer review system and led to a rapid expansion in the use of preprint servers to disseminate related findings. In gathering and analyzing a large database of preprint papers, we have found evidence of hasty scientific research communication and reduced gender balance in early COVID-19 preprint research. The trade-off between speed and meticulousness in reporting findings produced imprecise language, awkward syntax, and other infelicities. Our findings also revealed that greater participation of female authors tends to produce more readable papers. Such evidence attests the need for more gender balance and the necessity of moving from gender balance to gender equity, including in pay^81^ and career opportunities^81^. In sum, the quantitative analysis of preprint research from the early months of the COVID-19 pandemic has served to highlight the exacerbation of preexisting issues in the medical research environment. In order to communicate effectively with the public and policy makers, researchers must be cognizant of the readability of their work and explicitly communicate key takeaways and limitations. Paying attention to such aspects of science dissemination is crucial, given the current news ecosystems and the unprecedented pace at which information is shared – not only by researchers themselves but also by journalists who feature their preprints. Simultaneously, it is important to maintain confidence in the high quality of scientific work and to avoid compromising methodological rigor for the sake of readability. The scientific community must take this to heart and explore methods for fostering an inclusive, equitable, and responsive medical research system that is able to tackle global crises nimbly, while avoiding the pitfalls seen in the preprint ecosystem.

## Data Availability

All data are publicly available and a replication file will be made available on the Harvard Dataverse upon publication.

---

**Appendix Table 1:** Gender representation in preprint authorship by sub-discipline. For sub-disciplines with more than 1,000 preprint articles, the table presents the number of articles in that sub-discipline, the corresponding gender balance ratio, and the counts of male-authored-only and female-authored-only papers. The gender balance ratio is defined as the number of female authors over the total number of authors, so a value closer to 0.5 indicates balance in gender representation. The difference between COVID-19-related preprints and other research studies is similar to the difference between epidemiology (more male-dominated, with a gender balance ratio of 0.30) and cancer biology (less male-dominated, with a gender balance ratio of 0.35) preprints.

|  | Sub-discipline | Article Count | Gender Ratio | Male-only | Female-only |
| --- | --- | --- | --- | --- | --- |
| 1 | animal-behavior-and-cognition | 1313 | 0.31 | 468 | 104 |
| 2 | biochemistry | 2732 | 0.31 | 526 | 73 |
| 3 | bioengineering | 1831 | 0.28 | 507 | 80 |
| 4 | bioinformatics | 8199 | 0.24 | 3073 | 279 |
| 5 | biophysics | 3642 | 0.24 | 1505 | 372 |
| 6 | cancer-biology | 2929 | 0.35 | 396 | 86 |
| 7 | cell-biology | 4313 | 0.37 | 607 | 174 |
| 8 | developmental-biology | 2499 | 0.39 | 382 | 129 |
| 9 | ecology | 3699 | 0.30 | 1249 | 290 |
| 10 | epidemiology | 3170 | 0.30 | 965 | 127 |
| 11 | evolutionary-biology | 5326 | 0.27 | 1967 | 302 |
| 12 | genetics | 4473 | 0.32 | 883 | 142 |
| 13 | genomics | 5435 | 0.30 | 1043 | 125 |
| 14 | immunology | 2381 | 0.37 | 277 | 70 |
| 15 | infectious-diseases | 1539 | 0.32 | 276 | 22 |
| 16 | microbiology | 7277 | 0.37 | 1067 | 292 |
| 17 | molecular-biology | 2800 | 0.35 | 445 | 86 |
| 18 | neuroscience | 14954 | 0.31 | 4007 | 709 |
| 19 | physiology | 1146 | 0.34 | 225 | 58 |
| 20 | plant-biology | 2548 | 0.33 | 499 | 96 |
| 21 | systems-biology | 2246 | 0.25 | 781 | 75 |

**Appendix Figure 1:**
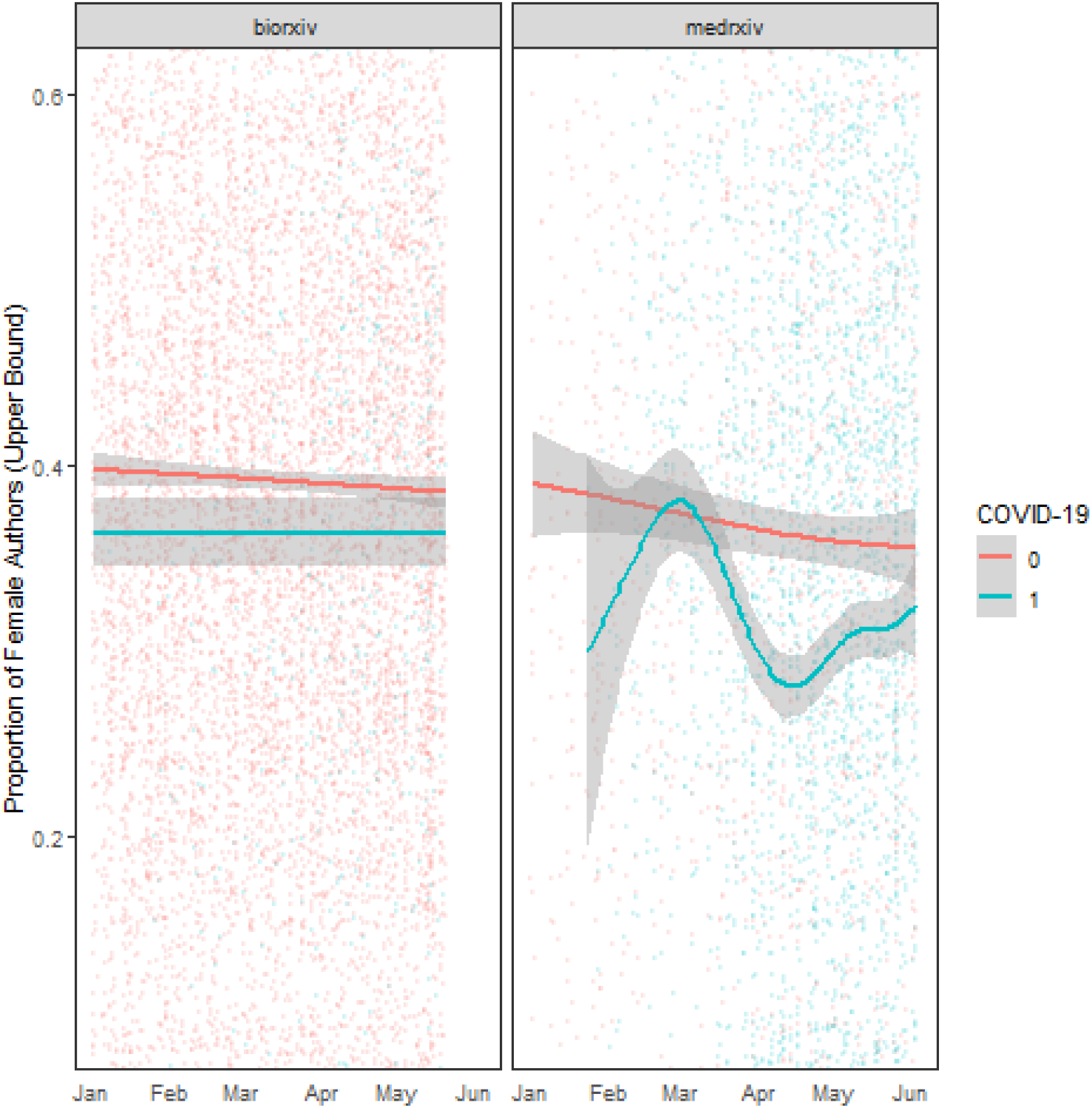
Female authorship among bioRxiv and medRxiv preprints over time, between January and June of 2020, when all indeterminate gender cases are imputed as female. On each panel, the y-axis represents the proportion of female authors. The lines correspond to daily averages (based on preprint release date, first version) among COVID-19-related papers (blue) and other research studies (pink), while shaded areas represent 95% confidence intervals. Each dot maps to a preprint article posted on either bioRxiv (left panel) or medRxiv (right panel). Of note, we still observe a sizable gender gap in COVID-19-related preprint authorship when considering the upper bound of the proportion of female authors.

